# Bivalent RSVpreF effectiveness against RSV-related lower respiratory tract disease hospitalization and ED visits across three RSV Seasons

**DOI:** 10.64898/2026.07.08.26357564

**Authors:** Sara Y. Tartof, Evan J. Zasowski, Negar Aliabadi, Gabriella Goodwin, Jeff Slezak, Vennis Hong, Timothy B. Frankland, Bradley Ackerson, Qing Liu, Sally Shaw, Sabrina Welsh, Banshri Kapadia, Brigitte C Spence, Gregg S. Davis, Joseph A. Lewnard, Hina Chowdhry, Michael Dutro, Erica Chilson, Alejandro Cane, Kyla Hayford, Elizabeth Begier

**Affiliations:** Department of Research & Evaluation, Kaiser Permanente Southern California, Pasadena, CA; Pfizer, Inc., New York, NY; Kaiser Permanente Hawaii Center for Integrated Health Care Research, Honolulu, HI; Southern California Permanente Medical Group, Harbor City, CA; Division of Epidemiology and Division of Infectious Diseases and Vaccinology, School of Public Health, and Center for Computational Biology, College of Engineering, University of California, Berkeley, Berkeley, CA

**Keywords:** respiratory syncytial virus, bivalent RSVpreF, lower respiratory tract disease, vaccine effectiveness, test-negative, hospitalization

## Abstract

**Introduction:** RSV vaccines reduce the risk of severe outcomes such as hospitalization and emergency department (ED) visits for at least 2 RSV seasons following vaccination. No data have been published on real-world RSV vaccine effectiveness (VE) beyond the second season after vaccination. This study evaluates bivalent RSVpreF VE against RSV-related lower respiratory tract disease (LRTD) hospitalizations/ED visits throughout 3 seasons after vaccination.

**Methods:** This retrospective test-negative case-control evaluates bivalent RSVpreF VE among adults aged ≥60 years at Kaiser Permanente Southern California with LRTD over 3 RSV seasons (11/24/2023–4/18/2026). Cases were RSV-positive without coinfection. Controls were negative for RSV, hMPV, influenza, SARS-CoV-2, and positive for a non-vaccine preventable disease pathogen. Exposure was bivalent RSVpreF (Abrysvo) receipt ≥21 days before LRTD. Adjusted VE was estimated using odds ratios from multivariable logistic regression or generalized estimating equations.

**Results:** Overall, adjusted VE against RSV-related LRTD hospitalization/ED visits was 80% (95% CI:68–87), 70% (95% CI:53–81), and 51% (95% CI:-12–78) in the first, second, and third season after vaccination, respectively. Among non-immunocompromised individuals, adjusted VE against RSV-related LRTD was 87% (95% CI:75–94), 76% (95% CI:55–87), and 58% (95% CI:-21–86) in the first, second, and third season after vaccination, respectively. Adjusted VE across the 3 combined seasons was 73% (95% CI: 64-80) overall and 80% (95% CI: 69-87) among non-immunocompromised individuals.

**Conclusion:** These results suggest Bivalent RSVpreF provides protection against RSV-related LRTD outcomes for at least three seasons after vaccination in this population of older adults with a prevalence of comorbidities. This suggests RSV vaccination results in substantial individual and public health benefit.

**Key Summary Points:** *Why carry out this study?:*

- RSV vaccines reduce the risk of severe outcomes such as hospitalization and emergency department (ED) visits for at least 2 RSV seasons after vaccination
- Real-world RSV vaccine effectiveness (VE) beyond the second season after vaccination is currently unknown
- This study evaluates bivalent RSVpreF VE against RSV-related lower respiratory tract disease (LRTD) hospitalizations/ED visits throughout 3 seasons after vaccination.

*What was learned from this study?:*

- Overall, VE against RSV-related LRTD hospitalization/ED visits was 80% (95% CI:68–87), 70% (95% CI:53–81), and 51% (95% CI:-12–78) in the first, second, and third season after vaccination, respectively, and 73% (95% CI: 64-80) across the 3 combined seasons.
- These results suggest Bivalent RSVpreF provides protection against RSV-related LRTD outcomes for at least three seasons after vaccination in this population of older adults with a prevalence of comorbidities.
- RSV vaccination results in substantial individual and public health benefit.

## Introduction

Severe respiratory illness due to respiratory syncytial virus (RSV) causes hundreds of thousands of hospitalizations and tens of thousands of deaths in older adults annually in high-income countries.^1,2^ RSV vaccines reduce the risk of severe outcomes such as hospitalization and emergency department (ED) visits for at least 2 RSV seasons following vaccination.^3-7^ No data have been published on real-world RSV vaccine effectiveness (VE) beyond the second season after vaccination. This study evaluates bivalent RSVpreF VE against RSV-related LRTD hospitalizations/ED visits throughout 3 seasons after vaccination.

## Methods

This is a retrospective test-negative case-control evaluating VE among adults aged ≥60 years at Kaiser Permanente Southern California over 3 RSV seasons (11/24/2023–4/18/2026). Events were defined as first hospitalization/ED visit in each season with ≥1 LRTD ICD-10 discharge code in any position and concurrent multiplex PCR respiratory specimen testing (Roche Diagnostics cobas eplex RP2) of a nasal/nasopharyngeal swab collected 14 days prior to 3 days after admission. Full inclusion and exclusion criteria have been detailed previously.^7^ As RSV testing is infrequent in practice among hospitalized adults with LRTD^8^, we supplemented standard-of-care (SOC) testing with study-initiated salvage and RSV testing of swabs collected for SARS-CoV-2 and/or influenza testing using the same PCR assay as SOC specimens. Cases were RSV-positive without coinfection. Controls were negative for RSV, hMPV, influenza, SARS-CoV-2, and positive for a non-vaccine preventable disease pathogen.^3^ Exposure was defined as bivalent RSVpreF (Abrysvo) receipt ≥21 days before LRTD admission date.^3^ Adjusted VE was estimated using odds ratios from multivariable logistic regression or generalized estimating equations in analyses where individuals contributed multiple LRTD events. Analyses were performed using SAS version 9.4 (SAS Institute). KPSC’s institutional review board approved this study and waived informed consent requirements.

(ClinicalTrials.gov ID: NCT06077968)

## Results

Of 6,813 LRTD events among individuals aged ≥60 years with LRTD hospitalization/ED visits at study sites during the study period, 3,131 (46.0%) met inclusion criteria, of which 2,206 (70.5%) were hospitalizations and 925 (29.5%) were ED visits. Overall, 637 (20.3%) were vaccinated with bivalent RSVpreF and 911 (29.1%) were RSV-positive cases. The median age was 78 years (IQR 70─85), median Charlson comorbidity index was 4 (IQR 2─7), and 22.8% were immunocompromised. Adjusted VE against RSV-related LRTD hospitalization/ED visits was 80% (95% CI:68–87), 70% (95% CI:53–81), and 51% (95% CI:-12–78) in the first, second, and third season after vaccination, respectively (**Figure 1**). Among non-immunocompromised individuals, adjusted VE against RSV-related LRTD was 87% (95% CI:75–94), 76% (95% CI:55–87), and 58% (95% CI:-21–86) in the first, second, and third season after vaccination, respectively (**Figure 1**).

**Figure 1.**
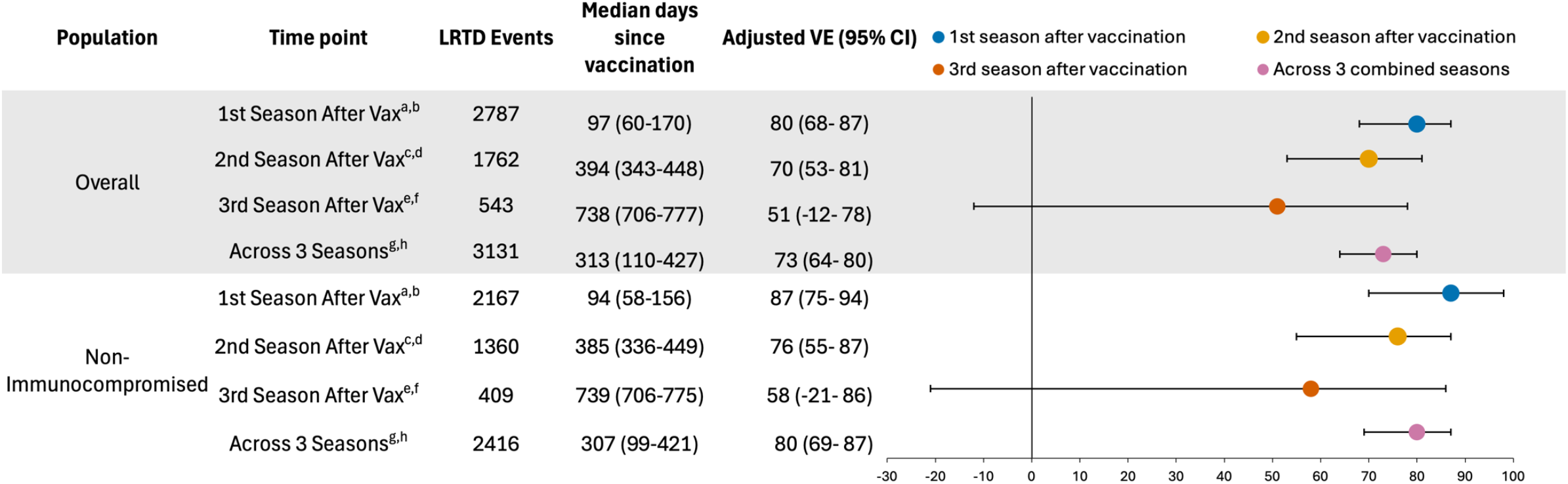
Bivalent RSVPreF Vaccine Effectiveness Estimates against RSV-related Lower Respiratory Tract Disease Hospitalization and Emergency Department Admissions Across 3 RSV Seasons. ^a^Includes those vaccinated from October, 2023 to March, 2024 and all unvaccinated, with LRTD events during the 2023-2024 RSV season, those vaccinated from April, 2024 to March, 2025 and all unvaccinated with LRTD events occurring during the 2024-2025 RSV season, and those vaccinated from April, 2025 to March, 2026 and all unvaccinated with LRTD events occurring during the 2025-2026 RSV season. ^b^Generalized estimating equation models for 1st season after vaccination were adjusted for age, sex, race/ethnicity, month of encounter, Charlson Index score, number of inpatient encounters in the year prior to index, flu vaccination, encounter facility, smoking status, diabetes status, and median household income. ^c^Includes those vaccinated from October, 2023 to March, 2024 and all unvaccinated, with LRTD events during the 2024-2025 RSV season and those vaccinated from April, 2024 to March, 2025 and all unvaccinated, with LRTD events during the 2025-2026 RSV season. ^d^Generalized estimating equation models for 2nd season after vaccination were adjusted for age, sex, race/ethnicity, month of encounter, Charlson Index score, BMI (body mass index), flu vaccination, encounter facility, diabetes status, and immunocompromised status. ^e^Includes those vaccinated from October, 2023 to March, 2024 and all unvaccinated, with LRTD events during the 2025-2026 RSV season. ^f^Logistic regression models for 3rd season after vaccination were adjusted for age, sex, race/ethnicity, month of encounter, Charlson Index score, flu vaccination, encounter facility, diabetes status, immunocompromised status, NDI (neighborhood deprivation index) quartile, number of virtual encounters in the year prior to index, and smoking status. ^g^Includes those vaccinated from October, 2023 to March, 2026 and all unvaccinated, with LRTD events during the 2023-2024, 2024-2025, and 2025-2026 RSV season. ^h^Generalized estimating equation models for across 3 seasons were adjusted for age, sex, race/ethnicity, month of encounter, Charlson Index score, flu vaccination, encounter facility, and diabetes status.

## Discussion

In this population of older adults with a high prevalence of comorbidities, our results suggest bivalent RSVpreF provides protection against RSV-related LRTD hospitalization and ED visits across three seasons after vaccination. These data are the first available real-world RSV VE for the third season after vaccination. Overall, VE waned modestly across three seasons with an absolute decrease of 10% in the first year and 19% in the second. A similar decline was observed among non-immunocompromised persons.

The only other available third season RSV vaccine effectiveness is from RSVpreF3’s randomized phase 3 trial, which showed 48% efficacy against symptomatic RSV-LRTD.^9^ These trial data are limited by exclusion of immunocompromised persons and lack of statistical power to assess events requiring hospitalization or ED visit.^9^ Our findings expand on this by examining bivalent RSVpreF, using a more severe outcome of RSV-related hospitalization/ED visit, and including a diverse real-world population at risk for severe disease, including those with immunocompromise.

Confidence intervals for third season after vaccination estimates were wide, and the small immunocompromised population’s estimate was not reported because it was too imprecise to allow meaningful interpretation. Further data are needed to more conclusively determine duration of protection amongst different populations, including immunocompromised individuals, to inform potential revaccination intervals. As the study is ongoing, this will be re-evaluated in future seasons.

Study strengths include minimal exposure misclassification with vaccination defined by electronic health record augmented by bi-directional data exchange with the California Immunization Registry, specimen salvage and RSV-testing to boost power and generalizability of RSV case detection, a multi-pathogen PCR to define a control group which minimizes bias, and a socio-demographically diverse study population. Limitations include lower RSV vaccine uptake in the first season after vaccine availability, decreasing number of available events for

each additional season post-vaccination limiting statistical power, reliance on data from a single health system, and possible residual confounding.

## Conclusion

Our results suggest Bivalent RSVpreF provides protection against RSV-related LRTD outcomes for at least three seasons after vaccination resulting in substantial individual and public health benefit.

## Data Availability

Deidentified participant data and protocol that support the findings of this study may be made available from the investigative team following publication in the following conditions: (1) agreement to collaborate with the study team on all publications, (2) provision of external funding for administrative and investigator time necessary for this collaboration, (3) demonstration that the external investigative team is qualified and has documented evidence of training for human subjects protections, and (4) agreement to abide by the terms outlined in data use agreements between institutions. Inquiries may be sent to Sara Tartof, PhD MPH at.

## Contributors

SYT, NA, EB, and EZ conceived this study. JMS, GG, TBF, and VH conducted the analysis. SYT, NA, JMS, GG, QL, and EB wrote the first draft of the protocol. SYT, NA, EZ, JMS and EB wrote the first draft of the manuscript. All authors contributed to the study design, drafting the protocol, and edited the manuscript for important intellectual content. All authors gave final approval of the version to be published. All authors had final responsibility for the decision to submit for publication.

VH, BK, GG, TBF and JMS had full access to all the data in the study and take responsibility for the integrity of the data and the accuracy of the data analysis.

## Ethics/Ethical approval

KPSC’s institutional review board approved this study and waived informed consent requirements.

